# Standardizing COVID-19 surveillance data into the OMOP common data model: a first implementation case study from Senegal

**DOI:** 10.64898/2026.07.01.26357078

**Authors:** Ousmane Diop, Rachel Odhiambo, Ousmane Diouf, Reipeinter Momanyi, Michael Ochola, Abdoulaye Samba Diallo, Abdou Padane, Steve Bycko Cygu, Miranda Barasa, Samuel Iddi, Agnes Kiragga, Moussa Sarr, Souleymane Mboup, Aminata Mboup

**Affiliations:** Data Management Department, Institut de Recherche en Santé, de Surveillance Épidémiologique et de Formation (IRESSEF), Dakar, Senegal; Data Science Program, African Population and Health Research Center (APHRC), Nairobi, Kenya; Data Synergy and Evaluation Unit, African Population and Health Research Center (APHRC), Nairobi, Kenya; Department of Statistics and Actuarial Science, University of Ghana, Accra, Ghana

## Abstract

The COVID-19 pandemic highlighted the need for interoperable health data infrastructures supporting reproducible observational research. The Observational Medical Outcomes Partnership Common Data Model (OMOP CDM) provides a widely adopted standard for harmonizing heterogeneous health data, but adoption remains limited in francophone Africa where language barriers and non-standardized surveillance systems pose additional challenges. We developed a complete Extract–Transform–Load (ETL) pipeline to convert a heterogeneous Senegalese COVID-19 surveillance dataset into OMOP CDM version 5.4. Source data recorded in French were translated into English through an iterative process interleaved with vocabulary mapping using ATHENA and Usagi. Semantic standardization used SNOMED CT for conditions, LOINC for measurements, and RxNorm for drugs. All 214 mappings underwent expert review by clinical and data science specialists. Data quality was assessed using the OHDSI Data Quality Dashboard (DQD) and Achilles. The standardized database achieved complete transformation (100%) for eight of the eleven source-populated domain tables, including person, visit_occurrence, measurement, and death. Partial transformation was observed for condition_occurrence (95.3%) and observation (68.1%), primarily due to incomplete vocabulary coverage for occupation categories and context-specific variables. The DQD produced an overall pass rate of 97% and a corrected pass rate of 98%, comparable to other published African OMOP implementations. Among the 19 data-quality failures, conformance and completeness issues predominated; the conformance failures were largely foreign-key checks, reflecting placeholder concept values (concept_id = 0) for metadata fields without meaningful equivalents in surveillance data. Iterative translation refinement was required when French-to-English translations did not align with OHDSI vocabulary terminology. This work documents, to our knowledge, the first OMOP CDM implementation on COVID-19 surveillance data in Senegal and francophone West Africa and provides a reusable methodological blueprint for future OMOP deployments in the region.

## Introduction

The growing digitization of health systems has led to an unprecedented increase in real-world health data from surveillance platforms, laboratory systems, and clinical care [1,2]. These data offer major opportunities for observational research and evidence-based public health decision-making. However, their scientific value is often limited by the lack of interoperability between heterogeneous data sources, particularly in low- and middle-income countries (LMICs) where information systems are frequently fragmented and non-standardized [3,4]. COVID-19 surveillance databases in such settings are a typical example of rich yet poorly interoperable data that are difficult to leverage for large-scale, reproducible analyses. [5]

To address this challenge, the Observational Medical Outcomes Partnership Common Data Model (OMOP CDM), developed within the Observational Health Data Sciences and Informatics (OHDSI) collaborative, has become one of the most widely used frameworks for large-scale observational health research worldwide [6,7]. OMOP provides a unified relational database structure and standardized vocabularies that support reproducible, multi-site analyses while preserving local data governance. It is embedded in a mature open-source analytical ecosystem including tools for data profiling (WhiteRabbit), vocabulary mapping (Usagi, ATHENA), data quality assessment (Data Quality Dashboard, Achilles), and cohort characterization (ATLAS) [7]. Among available health data standards, OMOP CDM was selected for this study for several reasons. Unlike Fast Healthcare Interoperability Resources (FHIR), which is primarily designed for real-time clinical data exchange between systems, OMOP CDM is specifically optimized for retrospective observational research and large-scale analytics [7]. Compared with i2b2, OMOP offers a more comprehensive and actively maintained analytical ecosystem with integrated quality assessment and characterization tools [8]. OMOP CDM has been successfully implemented in North America, Europe, and parts of Asia to support pharmacoepidemiology, vaccine safety monitoring, and real-world effectiveness studies [9,12–14]. In Africa, a growing number of implementations have been documented, though adoption remains limited. These include OMOP-based data sharing for African population health data [10] and the INSPIRE datahub, a pan-African integrated suite of services for harmonizing longitudinal population health data across multiple Health and Demographic Surveillance System (HDSS) sites [11], a COVID-19 sero-surveillance implementation in the Nairobi Urban HDSS in Kenya [15], an artificial intelligence-assisted harmonization project on SARS-CoV-2 data in Rwanda [16], and a recent OMOP CDM implementation for respiratory disease data at Douala General Hospital in Cameroon [17]. Broader regional networks such as ALPHA, INDEPTH, and the South African Population Research Infrastructure Network (SAPRIN) have likewise advanced the harmonization of longitudinal population health data in Africa [11].

Despite this progress, adoption in francophone West Africa remains limited. Additional barriers in this region include French-language clinical data that require translation before semantic mapping, limited access to standardized terminologies, and heterogeneous surveillance infrastructures [5]. To our knowledge, no fully documented OMOP CDM implementation on COVID-19 surveillance data has been reported in this subregion.

In Senegal, SARS-CoV-2 testing and surveillance during the pandemic involved multiple institutions, including public hospitals, diagnostic laboratories, and research centers, resulting in highly heterogeneous data sources collected under emergency operational conditions [18–20]. Standardizing these data into an international common data model offers strategic advantages: it enables reproducible epidemiological analyses, facilitates integration into global research networks, and creates a reusable digital infrastructure for future outbreaks and public health studies. At the same time, such a transformation raises important technical and semantic challenges related to terminology translation, mapping of local codes to international standards, and management of inconsistent real-world data [21].

In this study, we report the implementation of the OMOP Common Data Model for COVID-19 surveillance data in Senegal. Our objectives were threefold: (1) to develop and document a complete Extract–Transform–Load (ETL) pipeline converting a heterogeneous, multi-source SARS-CoV-2 testing registry into OMOP CDM version 5.4 using open-source OHDSI tools; (2) to assess the quality of the standardized database using the OHDSI Data Quality Dashboard and Achilles; and (3) to demonstrate the analytical utility of the standardized data through descriptive characterization using the ATLAS platform [22]. Beyond the COVID-19 context, this work is intended to provide a reusable methodological blueprint for future OMOP deployments in public health surveillance in West Africa.

## Materials and Methods

### Study design and data sources

This study is a retrospective observational data standardization study based on routine COVID-19 surveillance data collected in Senegal covering the period from 2020 to 2022 as part of the national public health response to the SARS-CoV-2 pandemic [18–20]. The source dataset was generated through national diagnostic and surveillance activities coordinated by public health authorities in collaboration with research institutes and authorized diagnostic laboratories. IRESSEF served as a central testing and research hub, performing RT-PCR diagnostics and collecting epidemiological data in collaboration with public health centers across the Dakar and Thies regions. Testing was conducted at IRESSEF’s own facilities as well as at partner sampling sites, covering both clinical referrals and community-based screening. A substantial proportion of the tested population consisted of international travelers screened at the Blaise Diagne International Airport (AIBD) as part of the national border health surveillance protocol, alongside symptomatic suspected cases, confirmed contacts, and individuals identified through community screening campaigns [18–20]. All collected data were centralized into a single relational flat file that served as the primary source for the Extract–Transform–Load (ETL) pipeline.

Before any transformation or analysis, all personally identifiable information was removed or replaced with anonymized surrogate identifiers in accordance with Senegalese Law No. 2008-12 of 25 January 2008 on the Protection of Personal Data [23]. A unique internal person_id was generated for each individual using deterministic matching on the national_id field, enabling longitudinal linkage across repeated tests and visits while preserving confidentiality.

The de-identified dataset was accessed for research purposes on January 1, 2023. The research team had access only to anonymized surrogate identifiers and at no point had access to information that could identify individual participants during or after data collection.

### Source data description

The source dataset consisted of a COVID-19 surveillance registry comprising 67 variables collected through routine diagnostic and epidemiological activities [18–20]. These variables captured sociodemographic characteristics (e.g., age, sex, region, marital status, occupation), clinical symptoms (e.g., fever, cough, headache, dyspnea), laboratory results (RT-PCR results), contextual information (sampling site, reason for testing, travel history), limited comorbidity data (asthma), treatment information (medication variables), and patient outcomes (alive/deceased). The overall structure of the source variables is summarized in Table 1.

**Table 1.**
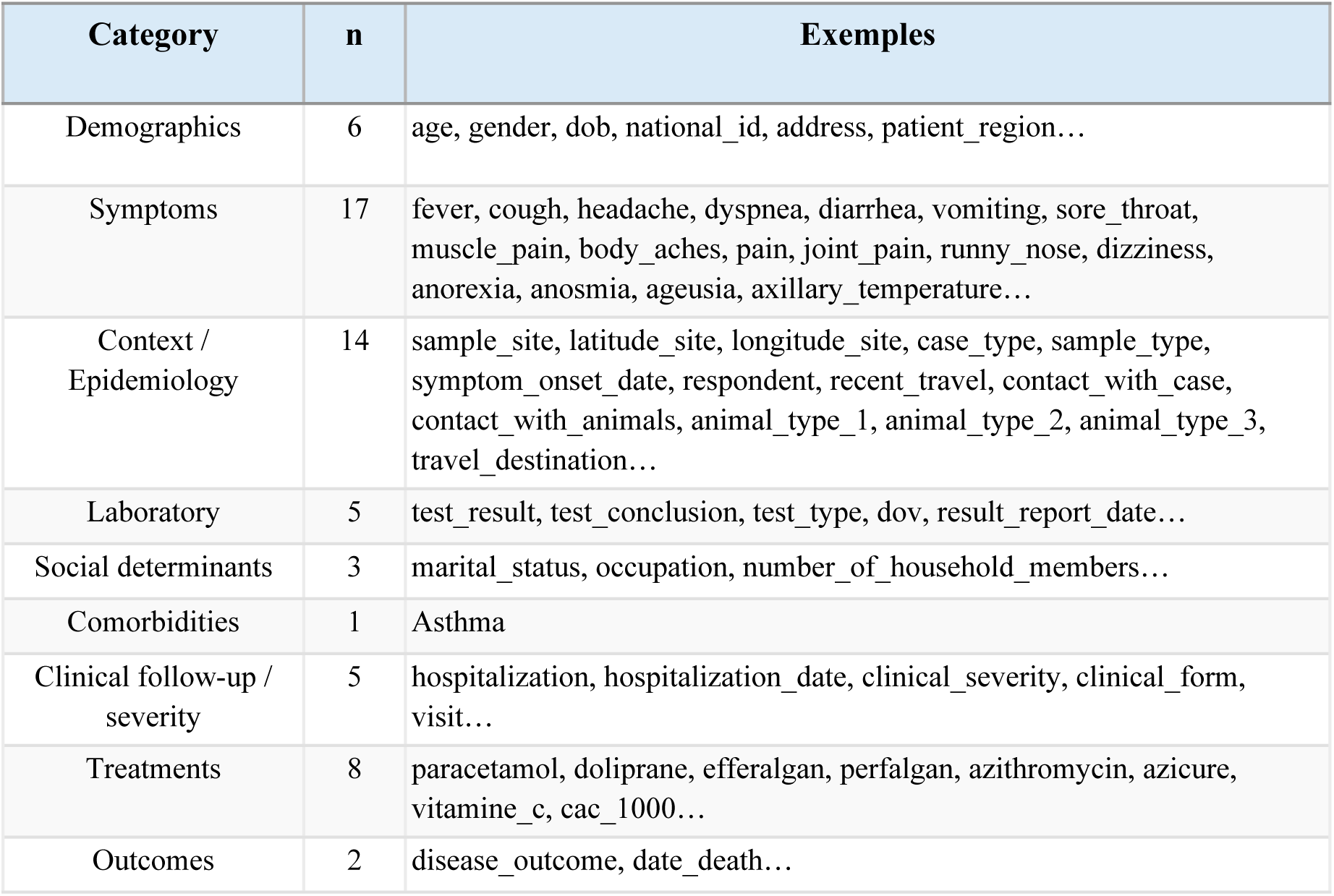
Representative variables per category.

A structured data dictionary including variable definitions, data types, distributions and proportions of missing data for all 67 source variables is provided in S1 Table. Core epidemiological and laboratory variables showed low levels of missingness, consistent with operational surveillance standards. The study population included 13,090 unique tested individuals and 13,406 testing visits, reflecting repeated testing for a subset of individuals.

### ETL pipeline and OMOP transformation

An Extract–Transform–Load (ETL) pipeline was developed to convert the heterogeneous COVID-19 surveillance dataset into the OMOP Common Data Model version 5.4 [6]. The pipeline followed the OHDSI methodological framework and was implemented using Python and SQL, with all scripts maintained under version control and publicly available at the study’s GitHub repository.

The structural design of the transformation was documented using the OHDSI tools WhiteRabbit and Rabbit-in-a-Hat [24], which enabled automated profiling of the source dataset and graphical documentation of the mappings between source variables and OMOP target tables (Fig 1, Fig 2). WhiteRabbit generated a scan report of the source file summarizing variable types, value distributions, and completeness, which guided the subsequent mapping decisions.

**Fig 1.**
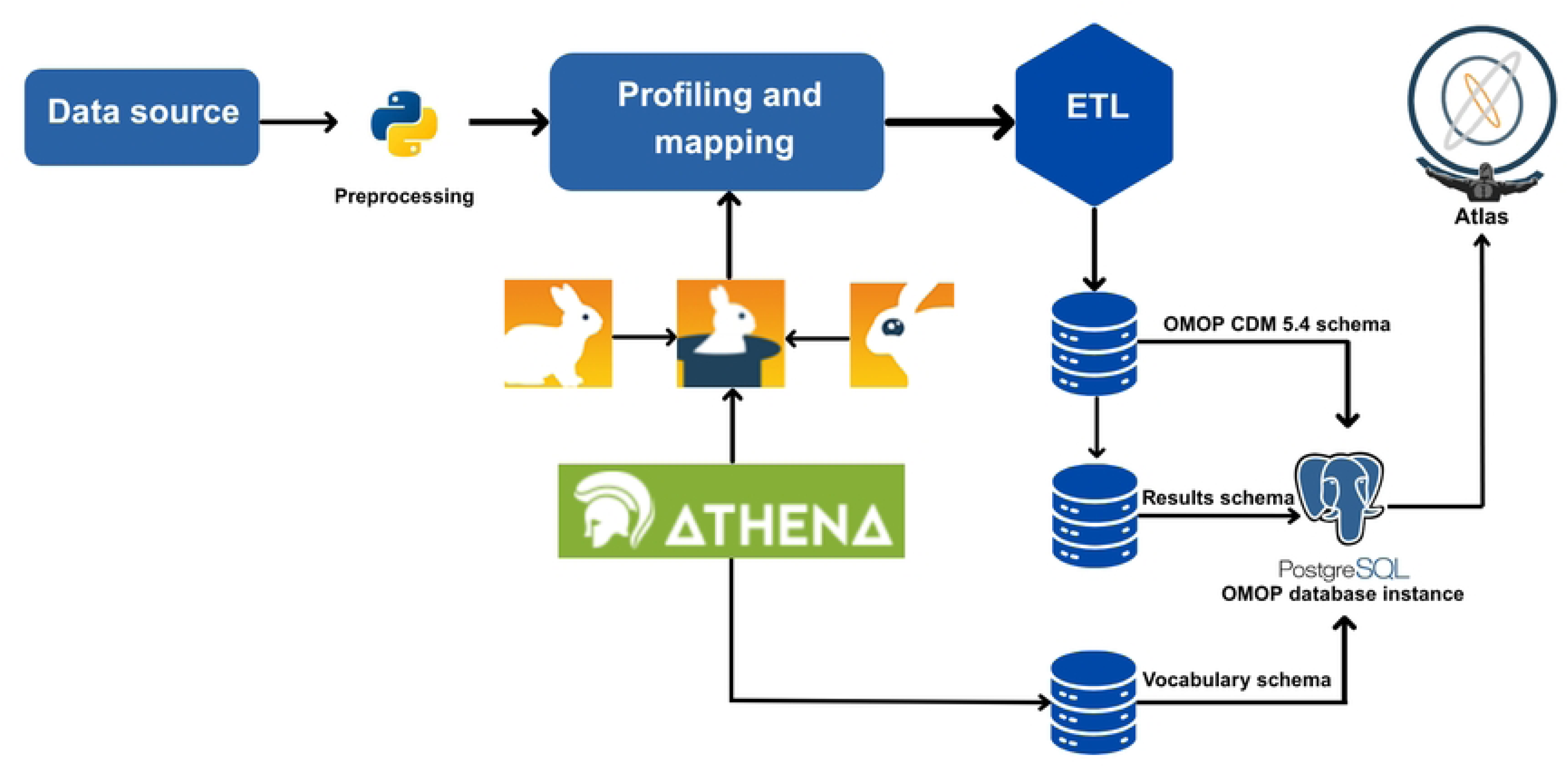
ETL Pipeline from source data to OMOP format.

**Fig 2.**
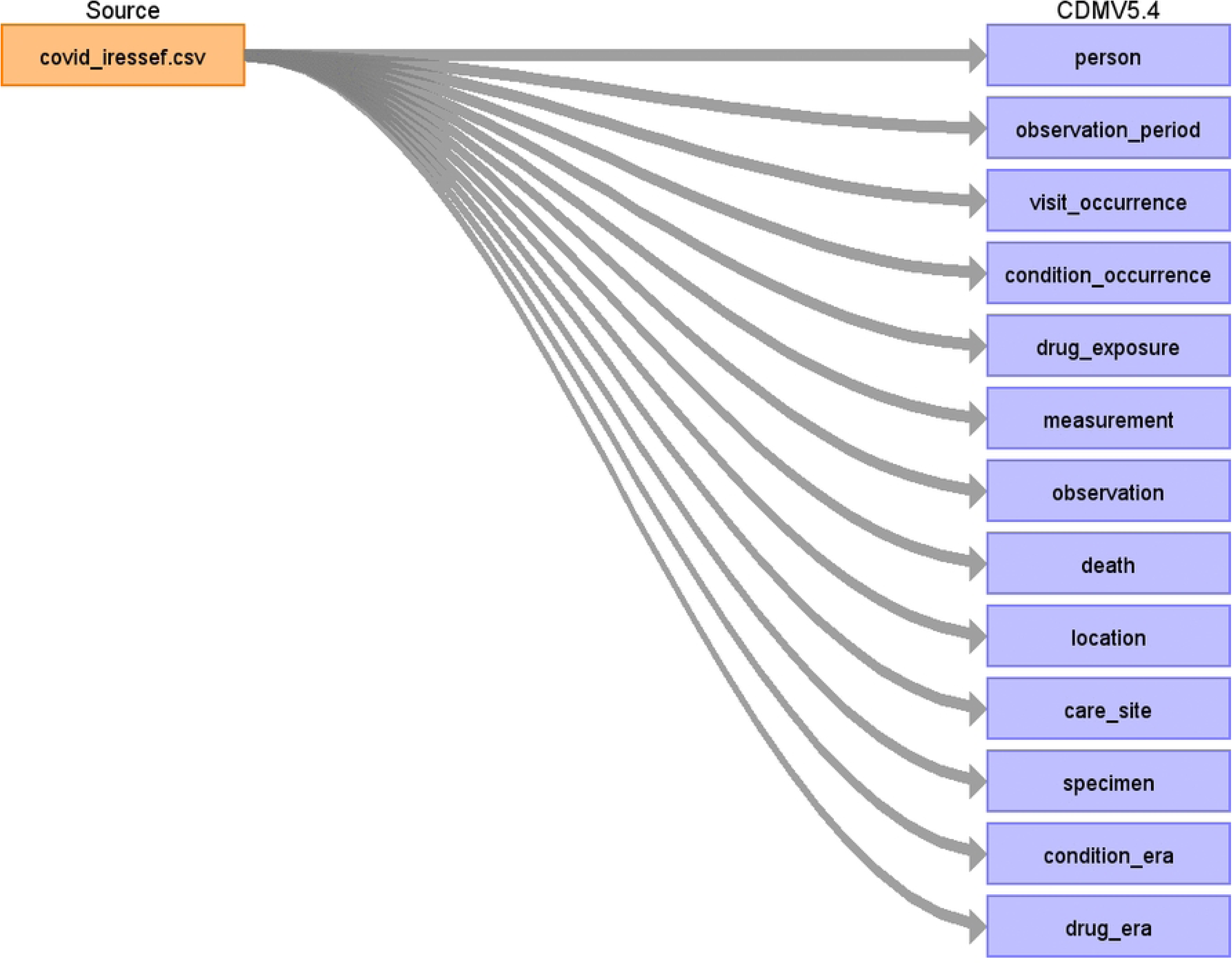
Mapping using Rabbit in a Hat.

During the extraction phase, the centralized flat source file was imported into a PostgreSQL processing environment. Structural validation checks were applied to verify variable formats, data types, value ranges, and internal consistency. A key preprocessing step involved the translation of all French-language source variables and categorical values into English, as the OHDSI vocabulary ecosystem operates exclusively with English-language terminology.

This translation was performed manually by the study team and covered variable names, categorical labels (e.g., “fièvre” → “fever”, “décédé” → “deceased”), and free-text values such as job titles (e.g., “commerçant” → “merchant”, “élevage” → “livestock farming”) and geographic entities. Semantic standardization was performed using OMOP standard vocabularies accessed through ATHENA [25]: SNOMED CT for clinical symptoms and conditions, LOINC for laboratory measurements and categorical values, RxNorm and RxNorm Extension for drug exposures, and Gender vocabulary for sex [7]. Semi-automated concept matching was performed using Usagi [26], which generates candidate mappings based on string similarity between source terms and OHDSI vocabulary concepts.

During the Usagi matching process, it became apparent that several initial English translations did not align well with the terminology used in OHDSI vocabularies, resulting in low match scores or incorrect candidate concepts. This required an iterative refinement approach: when Usagi returned poor matches for a translated term, the translation itself was revised to better approximate the vocabulary used in SNOMED CT, LOINC, or RxNorm before re-running the matching. For example, the French term “courbatures” was initially translated as “body aches”, which yielded a low Usagi match score (0.23) and was ultimately mapped to “Soreness” (SNOMED concept 4325423) after expert review. Similarly, “mal de gorge” was translated as “sore_throat” and matched to “Has a sore throat” (concept 4036632, score 0.36). These low scores reflect the semantic distance between translated French clinical terms and the English-language OHDSI vocabulary, rather than incorrect mappings.

Local drug brand names posed additional challenges. The surveillance data recorded commercial names commonly used in Senegal and francophone Africa (e.g., “Doliprane”, “Efferalgan” and “Perfalgan” for acetaminophen formulations; “Azicure” for azithromycin). Doliprane and Efferalgan were mapped to their corresponding branded drug forms in RxNorm Extension (concept IDs 43212028 and 43146071). During expert review, the candidate RxNorm Extension concept automatically suggested for “Perfalgan” was found to be incorrectly labelled (it referenced an aspirin formulation), and no correct branded concept was available for “Azicure”; these two products were therefore mapped to the corresponding standard ingredients in RxNorm - acetaminophen (1125315) and azithromycin (1734104), respectively. The vitamin C products (“vitamine_c” and “CAC 1000”) were mapped to the ascorbic acid ingredient (19011773). This step illustrates that automated string-similarity matching can return clinically incorrect candidate concepts, which must be verified by domain experts before loading. All mappings were manually reviewed and validated by clinical and data science experts. Although the source registry comprised 67 variables, semantic mapping operates at the level of distinct source values rather than variables: the 214 mappings correspond to the unique categorical values requiring a concept (for example, 103 occupation categories, 11 regions, marital-status categories, symptom terms, drug brand names and test-result categories), not to the 67 variables themselves.

Across all 214 mapped source values, the median Usagi match score was 0.85 (interquartile range: 0.67–1.00); 164 mappings (76.6%) were classified as APPROVED after expert review, while 50 (23.4%) remained FLAGGED, primarily corresponding to job title and geographic entities for which exact OHDSI vocabulary equivalents were unavailable.

The key semantic mappings between source variables and OMOP standard concepts are presented in Table 2. The complete mapping file is available in the study’s GitHub repository.

**Table 2.**
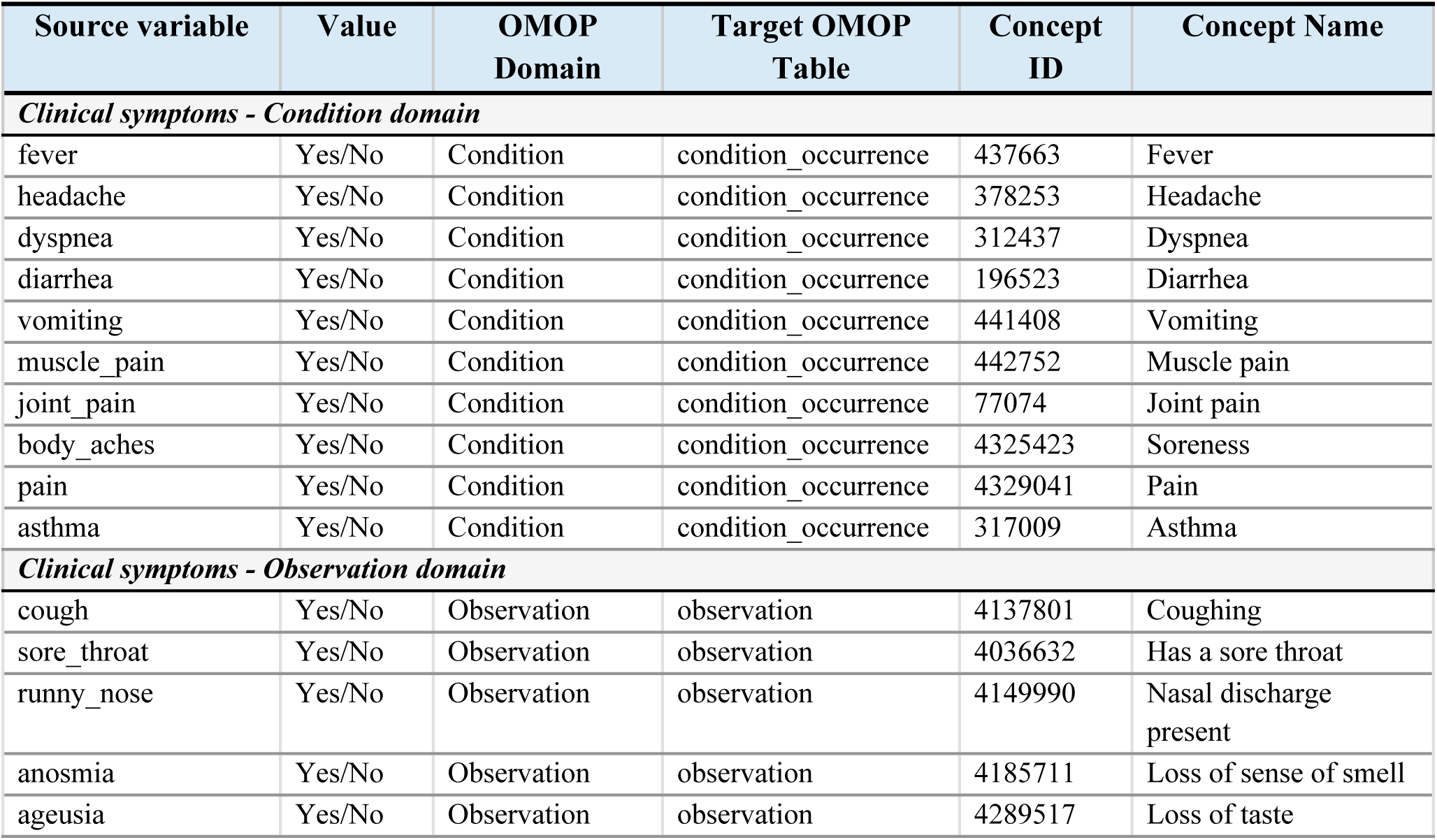
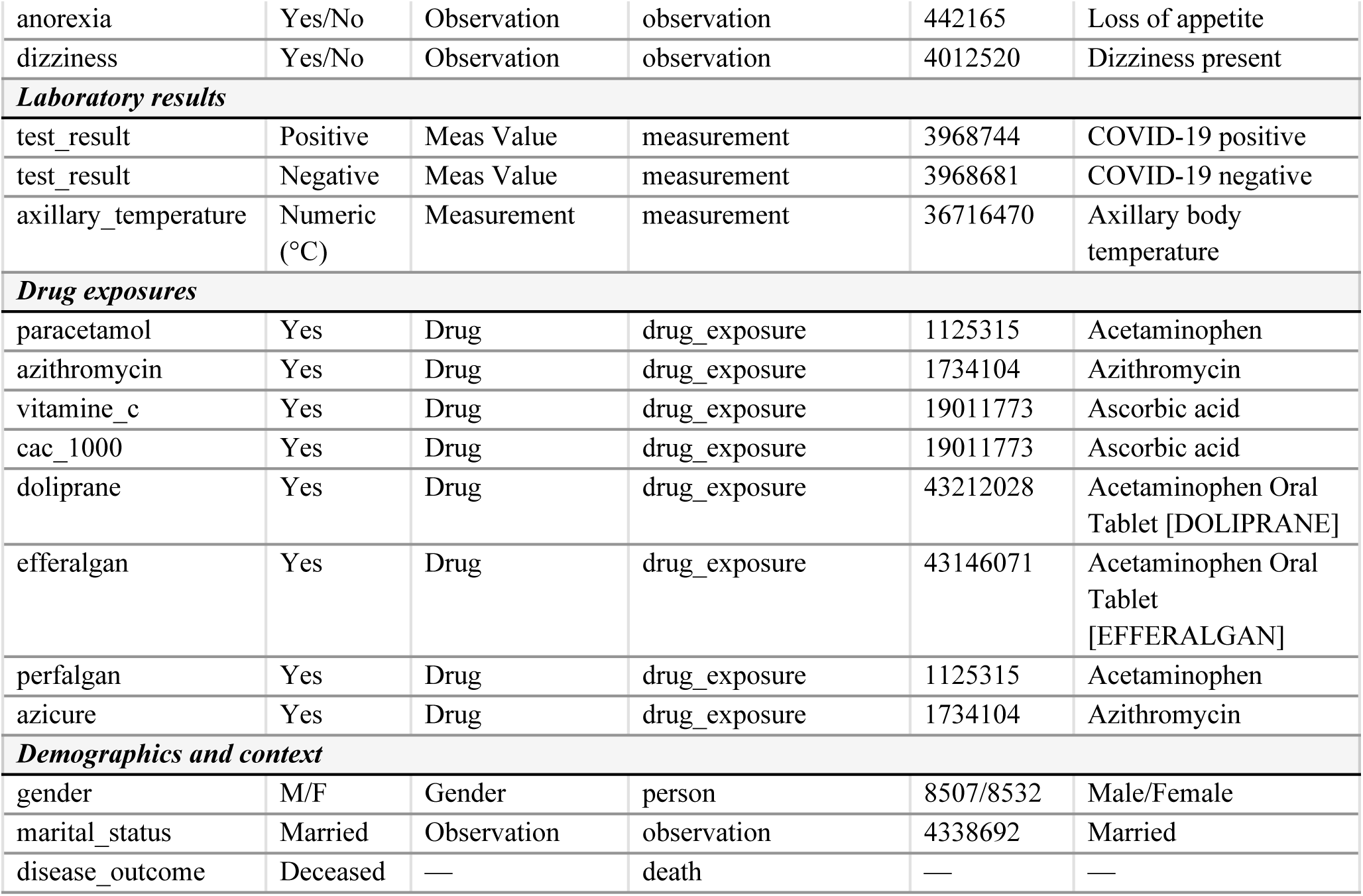
Semantic mapping of key source variables.

The transformation phase included data cleaning, normalization of categorical labels, harmonization of date formats, and management of missing or inconsistent values. Source variables were assigned to OMOP domain tables based on their semantic mapping in Usagi: clinical symptoms mapped to SNOMED Condition domain concepts were loaded into the condition_occurrence table, while symptoms mapped to SNOMED Observation domain concepts (e.g., coughing, nasal discharge) were loaded into the observation table.

Binary symptom indicators recorded both presence (“Yes”) and absence (“No”). Following OMOP conventions, only positive indicators (symptom = “Yes”) were transformed into condition_occurrence or observation records. Negative indicators were not recorded as separate observations, consistent with the OMOP principle that the absence of a record implies the absence of the condition. This design choice preserves analytical consistency with other OMOP databases but means that explicit negative symptom documentation is not retained in the standardized database.

Laboratory RT-PCR test results were transformed into standardized measurement records. Each testing visit generated up to two measurement records: one for the qualitative test result (positive, negative, doubtful or inconclusive) recorded as value_as_concept_id using standard SNOMED result concepts, and one for the axillary temperature when available. Serology results were handled separately using the same measurement structure. The RT-PCR testing procedure itself was not loaded into the procedure_occurrence table, as the measurement records already captured the complete testing information; the procedure_occurrence table was therefore left empty by design.

For the observation_period table, a record was created for each testing visit rather than one record per person, with the observation period start and end dates both set to the visit date. This design decision reflects the surveillance context in which only testing visit dates were available and no longitudinal follow-up end dates were recorded.

Contextual and sociodemographic variables, including marital status, job title, case type, disease severity, hospitalization status, contact history, and travel history, were loaded into the observation table. Geographic information was structured in the location table (11 regions) and care_site table (22 sampling facilities). The complete set of 103 job title categories was mapped to SNOMED Social Context concepts in the observation table, though not all source values had exact OHDSI vocabulary equivalents, resulting in partial mapping coverage for this variable. The loading phase consisted of populating a dedicated PostgreSQL database configured with the official OMOP CDM v5.4 schema. Standard bulk-loading procedures were used to insert standardized records into the main domain tables, including person, visit_occurrence, measurement, condition_occurrence, observation, drug_exposure, death, location, care_site, observation_period, and specimen. A specimen record was created for each RT-PCR testing visit to document the biological sample underlying each laboratory measurement. Referential integrity was systematically verified through primary and foreign key constraints. Each ETL execution generated detailed processing logs documenting record counts and data flows at each transformation stage, ensuring full traceability and reproducibility. The mapping of the person table is illustrated in Fig 3.

**Fig 3.**
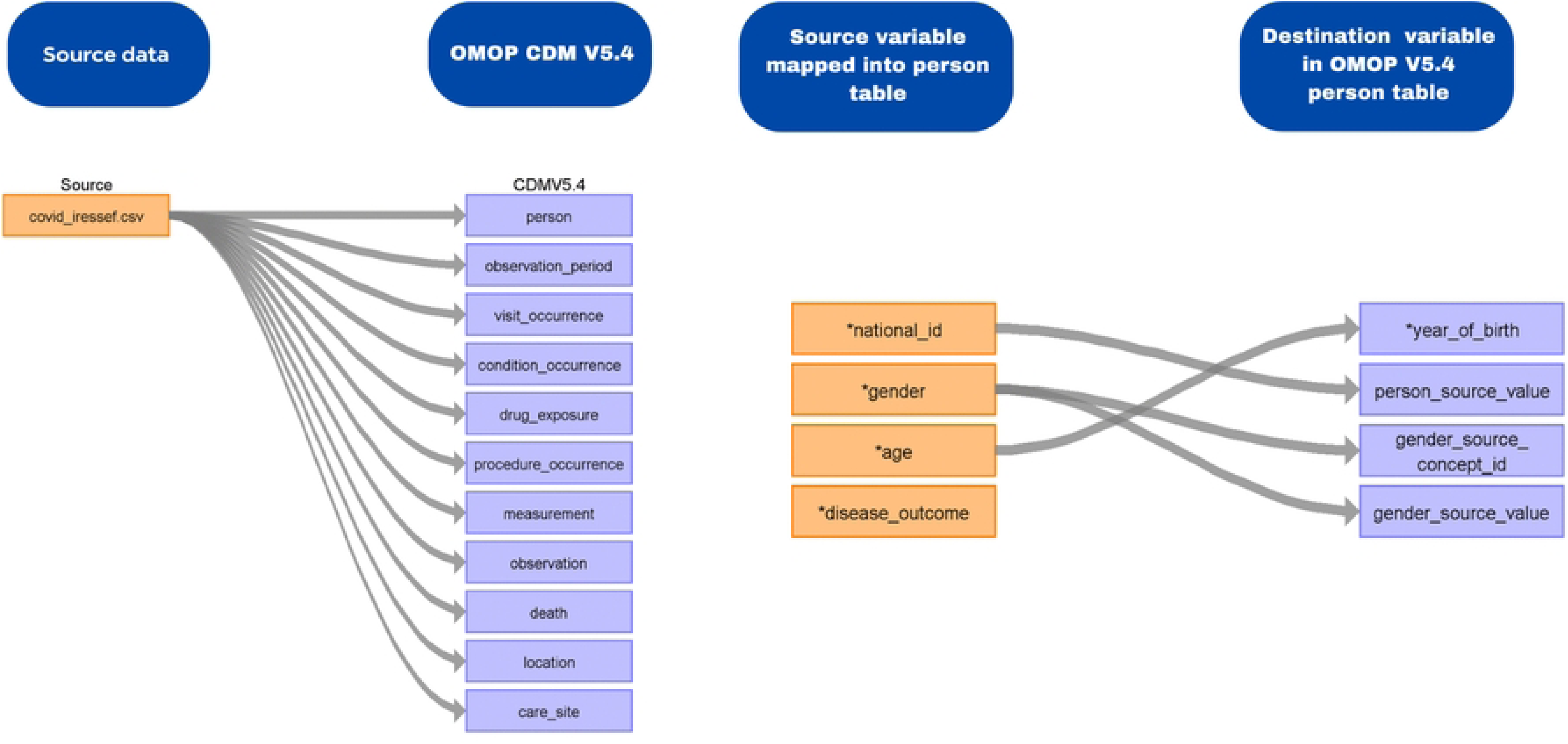
Mapping of person table into the OMOP CDM v5.4.

### Data quality assessment and descriptive characterization

After data loading, a comprehensive data quality assessment was conducted using the OHDSI Data Quality Dashboard (DQD) version 2.6.1 [27], which evaluates three dimensions of data quality: conformance (adherence to structural and syntactic standards), completeness (presence of expected values), and plausibility (logical consistency of recorded values).

The DQD executed a total of 2,312 checks across all populated OMOP tables. Following OHDSI recommendations [7], checks related to empty tables or unpopulated fields were classified as not applicable and excluded from the corrected pass rate. A detailed analysis of failed checks was performed to identify their root causes and assess their implications for the standardized database. Achilles was used for additional automated data characterization and structural validation [28]. Following successful data quality validation, descriptive cohort exploration and characterization were performed using the OHDSI ATLAS platform [22]. ATLAS was directly connected to the validated OMOP database and used to generate standardized descriptive indicators from the person, condition_occurrence, measurement, visit_occurrence, and death tables. ATLAS analyses were conducted using the default configuration settings without custom post-processing. ATLAS was used exclusively for exploratory and descriptive purposes, including the generation of population distributions, symptom prevalence, and mortality-related summaries. No predictive modeling, causal inference, or risk estimation analyses were performed at this stage.

## Results

The COVID-19 surveillance dataset was successfully transformed and loaded into a fully compliant OMOP Common Data Model version 5.4 PostgreSQL database following the ETL pipeline described in the Methods. Table 3 presents the record counts in the source data and the standardized CDM for each OMOP domain table, along with the transformation rate.

**Table 3.** Comparison between source data and standardized OMOP CDM database.

| OMOP CDM Table | Source records | CDM records | Transformation rate | Notes |
| --- | --- | --- | --- | --- |
| person | 13,090 | 13,090 | 100% | All unique individuals transferred |
| visit_occurrence | 13,406 | 13,406 | 100% | One record per testing visit |
| observation_period | 13,406 | 13,406 | 100% | One record per visit; start date = end date |
| measurement | 13,494 | 13,494 | 100% | RT-PCR qualitative results + axillary temperature |
| condition_occurrence | 11,846 | 11,290 | 95.3% | 556 records not loaded due to unmapped symptom values |
| observation | 71,915 | 48,995 | 68.1% | Partial mapping coverage for occupation and contextual variables |
| drug_exposure | 699 | 806 | 115.3% | One-to-many mapping of branded drug formulations |
| death | 52 | 52 | 100% | All death records transferred |
| location | 11 | 11 | 100% | Administrative regions |
| care_site | 22 | 22 | 100% | Sampling facilities |
| procedure_occurrence | 0 | 0 | N/A | Not populated; testing information captured in measurement |
| condition_era | - | 21,935 | N/A | Derived table generated from condition_occurrence using standard OHDSI era-construction logic; not transformed directly from source |
| drug_era | - | 803 | N/A | Derived table generated from drug_exposure using standard OHDSI era-construction logic; not transformed directly from source |
| specimen | 13,406 | 13,406 | 100% | One specimen record per testing visit (RT-PCR sample) |

Eight of the eleven OMOP tables populated directly from source data achieved a 100% transformation rate, indicating complete transfer of the corresponding source records. The condition_occurrence table had a transformation rate of 95.3%, with 556 records (4.7%) not loaded due to source symptom values that could not be mapped to standard OMOP concepts. The observation table showed the largest gap, with a transformation rate of 68.1%; this was primarily due to incomplete mapping coverage for the 103 job title categories recorded in the source data, many of which lacked exact equivalents in the OHDSI SNOMED vocabulary, as well as partial coverage for other contextual variables (animal contact type, country of provenance). The drug_exposure table count differed from the aggregate source count because several brand-name variables mapped to standard drug concepts at different levels of the RxNorm hierarchy: paracetamol and Perfalgan were mapped to the acetaminophen ingredient, while Doliprane and Efferalgan were mapped to distinct acetaminophen branded drug forms, so that a single individual could contribute several acetaminophen-related drug_exposure records.

The Data Quality Dashboard v2.6.1 executed a total of 2,312 checks across all populated OMOP tables. Of these, 1,295 (56.0%) were classified as not applicable because they targeted empty tables or unpopulated fields, 959 (41.5%) passed, 19 (0.8%) failed, and 39 (1.7%) returned SQL execution errors. Following OHDSI recommendations, not applicable checks and SQL errors were excluded from the corrected pass rate. The dashboard-reported overall pass rate, which counts not applicable checks as passed, was 97% (2,254/2,312); the corrected pass rate, restricted to applicable and executable checks, was 98% (959/978) (Fig 4).

**Fig 4.**
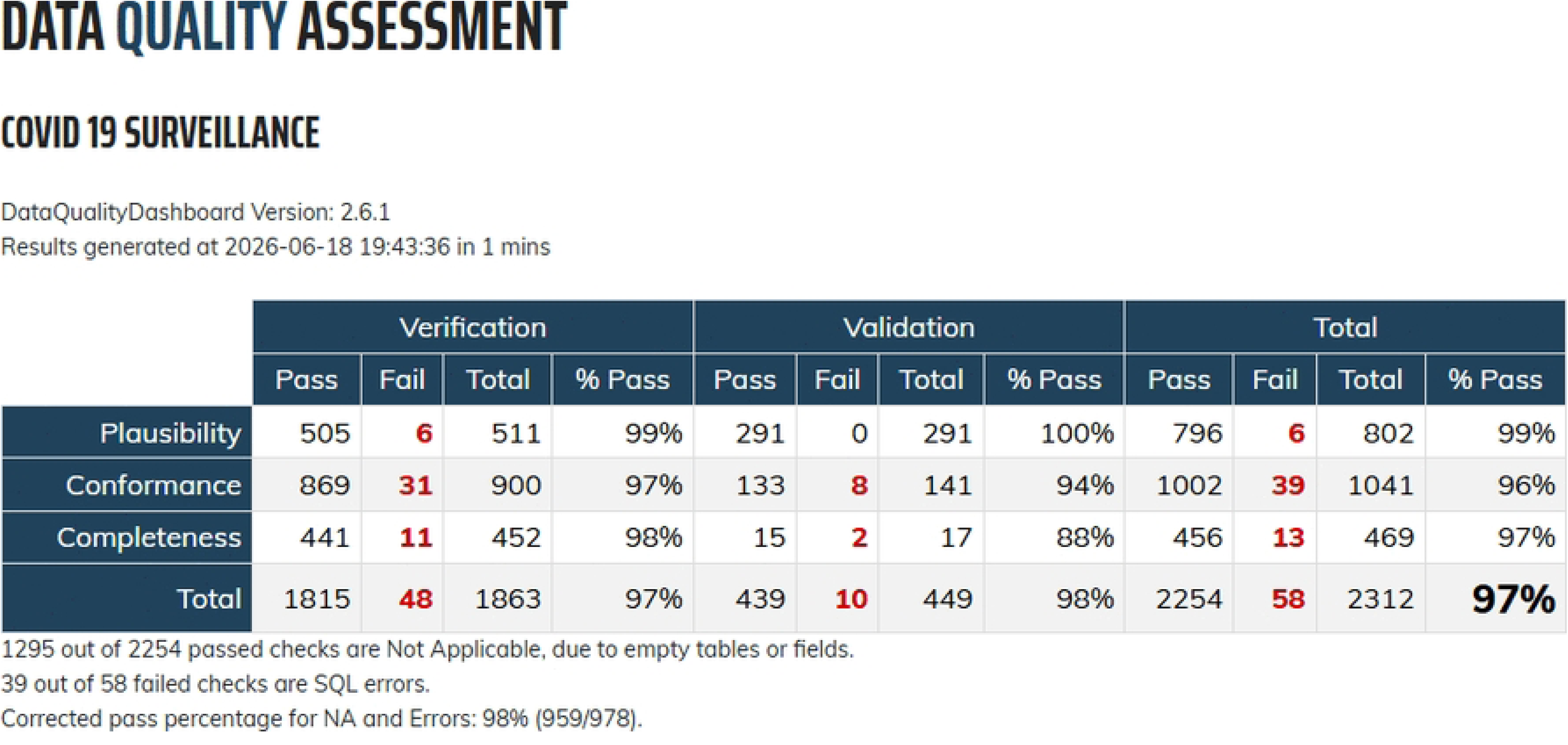
Results of the Data quality assessment.

Among the 19 failed checks, 8 were conformance failures, 9 were completeness failures, and 2 were plausibility failures (Table 4). The conformance failures were dominated by foreign-key checks (isForeignKey, fkDomain, fkClass; 7 of the 19 failures), reflecting the use of placeholder concept_id values (concept_id = 0) in type_concept_id and source_concept_id fields for which no meaningful OMOP vocabulary equivalent existed in the surveillance data - a pattern commonly reported for OMOP implementations based on non-clinical data sources [10,11,15]. The completeness failures concerned mainly the drug-related tables (drug_exposure, drug_era) and source/standard-concept fields, consistent with the limited and selective recording of treatment data in the surveillance registry (only ∼4.4% of individuals had a drug record). The 39 SQL errors were execution errors rather than data-quality failures: 35 concerned the COHORT and COHORT_DEFINITION result-schema tables (populated only when cohort definitions are executed in ATLAS, not during the ETL) and 4 concerned the VISIT_DETAIL and VISIT_OCCURRENCE tables; all were excluded from the corrected pass rate. Overall, these patterns are consistent with the known structural limitations of surveillance data and do not indicate systematic errors in the ETL pipeline.

**Table 4.** Summary of failed Data Quality Dashboard checks by category.

| DQD Category | Check type | N failed | Tables affected | Explanation |
| --- | --- | --- | --- | --- |
| Conformance | isForeignKey | 4 | condition_occurrence, observation, person | Foreign-key references to concept_id values not present in the CONCEPT table, predominantly concept_id = 0 placeholders used in type_concept_id and source_concept_id fields with no meaningful OMOP vocabulary equivalent in the surveillance data. |
| Conformance | fkDomain | 2 | condition_era, measurement | Records whose concept is assigned to a domain different from the one expected for the field, arising where source variables map to concepts spanning the Condition and Observation domains. |
| Conformance | fkClass | 1 | drug_era | Records whose concept class does not match the class expected for the field, in the derived drug_era table. |
| Conformance | isStandardValidConcept | 1 | observation | A negligible fraction of records referencing a non-standard or invalid concept. |
| Completeness | standardConceptRecordCompleteness | 3 | drug_exposure, drug_era, specimen | Proportion of records with standard_concept_id = 0 above the 5% threshold, for source values without an available standard concept. |
| Completeness | sourceConceptRecordCompleteness | 2 | death, drug_exposure | Source-concept fields left at 0 because source codes were mapped directly to standard concepts. |
| Completeness | sourceValueCompleteness | 2 | death, drug_exposure | Source-value fields with missing or unmapped values. |
| Completeness | measurePersonCompleteness | 2 | drug_exposure, drug_era | Low person-level coverage of the drug tables (~4.4% of individuals have a drug record), consistent with the selective recording of treatment data in the surveillance registry. |
| Plausibility | plausibleTemporalAfter | 1 | cdm_source | A temporal-ordering plausibility check on a metadata field of the cdm_source table. |
| Plausibility | plausibleValueLow | 1 | specimen | Values below the plausible range threshold in the specimen table. |

Table 5 describes the sociodemographic composition of the study population according to SARS-CoV-2 RT-PCR test result. Because the test result is the study outcome rather than a fixed design stratum, two complementary views are presented: Table 5 reports the composition of each result group as column percentages, while Table 6 reports the positivity proportion within each sociodemographic group as row percentages.

**Table 5.** Sociodemographic characteristics of the study population by SARS-CoV-2 RT-PCR test result.

| Characteristic | Negative<sup>1</sup> | Positive<sup>1</sup> | Doubtful<sup>1</sup> | Inconclusive<sup>1</sup> | Overall<sup>1</sup> |
| --- | --- | --- | --- | --- | --- |
| **Age categories** |  |  |  |  |  |
| 0-14 years | 380 (4.6%) | 127 (2.6%) | 8 (2.3%) | 4 (13%) | 519 (3.9%) |
| 15-24 years | 1,130 (14%) | 551 (11%) | 33 (9.6%) | 4 (13%) | 1,718 (13%) |
| 25-64 years | 5,890 (72%) | 3,570 (74%) | 261 (76%) | 17 (57%) | 9,738 (73%) |
| ≥65 years | 781 (9.5%) | 602 (12%) | 43 (12%) | 5 (17%) | 1,431 (11%) |
| **Gender** |  |  |  |  |  |
| Male | 4,002 (49%) | 2,432 (50%) | 166 (48%) | 15 (50%) | 6,615 (49%) |
| Female | 4,179 (51%) | 2,418 (50%) | 179 (52%) | 15 (50%) | 6,791 (51%) |
| **Location** |  |  |  |  |  |
| Dakar | 4,995 (61%) | 3,081 (64%) | 226 (66%) | 2 (6.7%) | 8,304 (62%) |
| Thies | 3,173 (39%) | 1,765 (36%) | 117 (34%) | 28 (93%) | 5,083 (38%) |
| Other region | 13 (0.2%) | 4 (<0.1%) | 2 (0.6%) | 0 (0%) | 19 (0.1%) |
| **Marital status** |  |  |  |  |  |
| Married | 5,630 (70%) | 3,492 (73%) | 267 (77%) | 11 (38%) | 9,400 (71%) |
| Single | 2,193 (27%) | 1,134 (24%) | 63 (18%) | 15 (52%) | 3,405 (26%) |
| Divorced | 118 (1.5%) | 62 (1.3%) | 4 (1.2%) | 0 (0%) | 184 (1.4%) |
| Widowed | 154 (1.9%) | 127 (2.6%) | 11 (3.2%) | 3 (10%) | 295 (2.2%) |
| Engaged | 0 (0%) | 1 (<0.1%) | 0 (0%) | 0 (0%) | 1 (<0.1%) |

<sup>1</sup> n (column %). Percentages are computed within each test-result category.

**Table 6.**
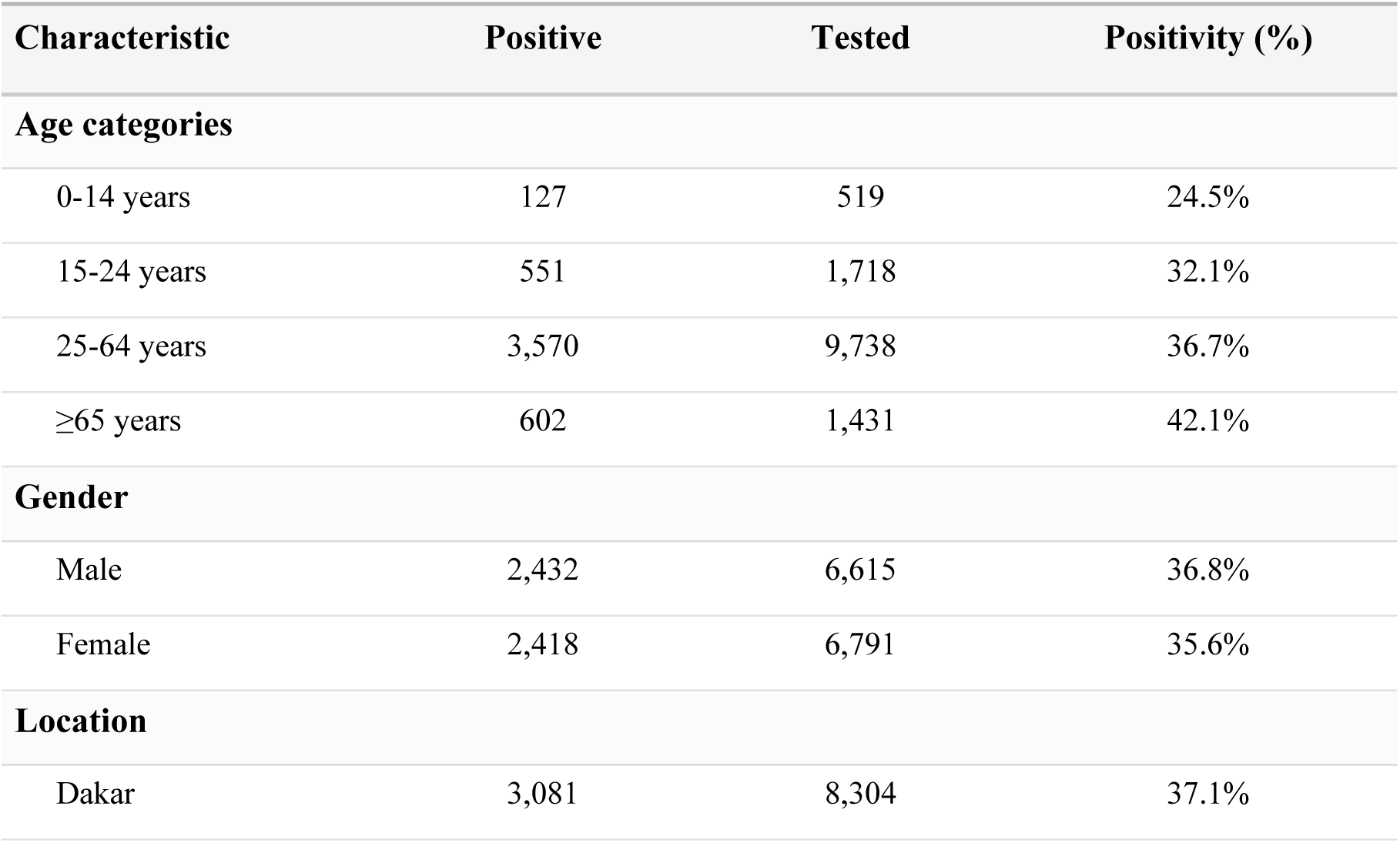
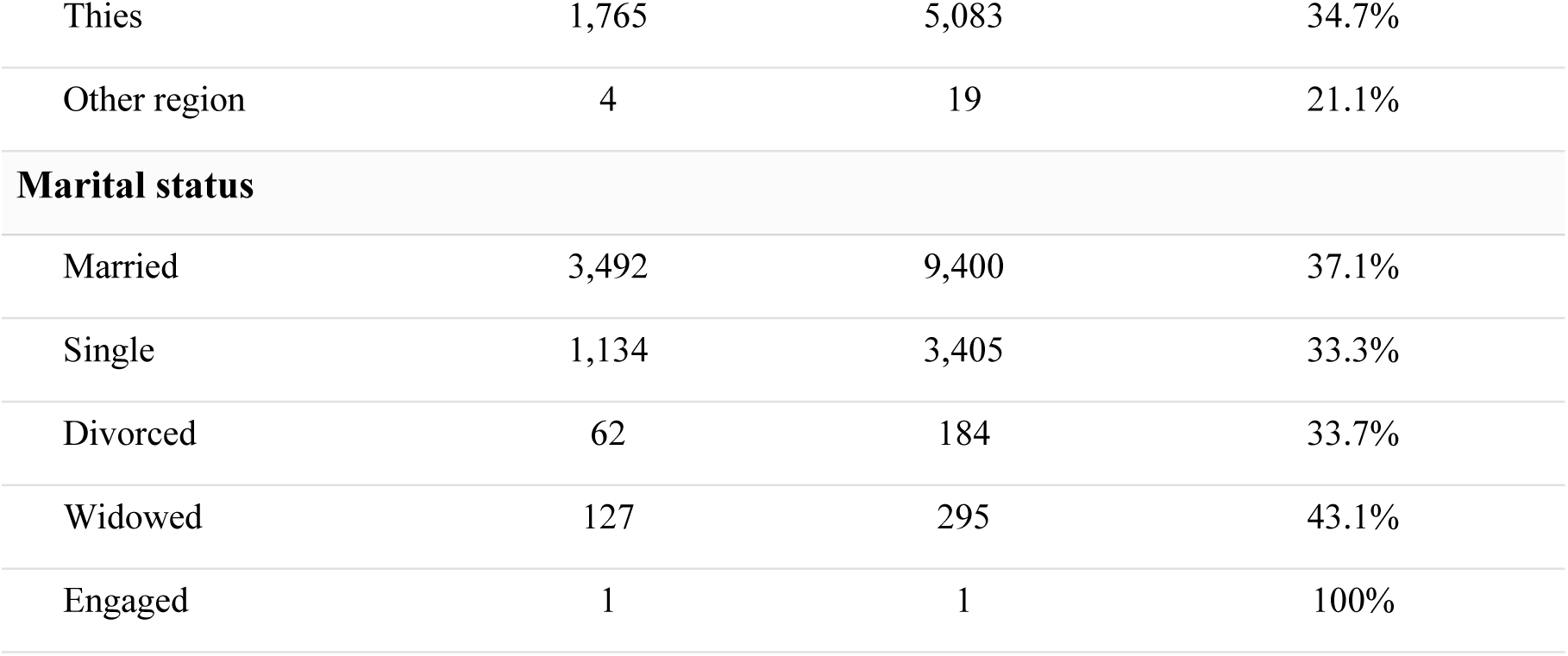
SARS-CoV-2 positivity proportion by sociodemographic characteristics (row percentages)

| Characteristic | Positive | Tested | Positivity (%) |
| --- | --- | --- | --- |
| **Age categories** |  |  |  |
| 0-14 years | 127 | 519 | 24.5% |
| 15-24 years | 551 | 1,718 | 32.1% |
| 25-64 years | 3,570 | 9,738 | 36.7% |
| ≥65 years | 602 | 1,431 | 42.1% |
| **Gender** |  |  |  |
| Male | 2,432 | 6,615 | 36.8% |
| Female | 2,418 | 6,791 | 35.6% |
| **Location** |  |  |  |
| Dakar | 3,081 | 8,304 | 37.1% |
| Thies | 1,765 | 5,083 | 34.7% |
| Other region | 4 | 19 | 21.1% |
| **Marital status** |  |  |  |
| Married | 3,492 | 9,400 | 37.1% |
| Single | 1,134 | 3,405 | 33.3% |
| Divorced | 62 | 184 | 33.7% |
| Widowed | 127 | 295 | 43.1% |
| Engaged | 1 | 1 | 100% |

The study population showed a balanced distribution by gender (50.7% female, 49.3% male), was predominantly composed of adults aged 25-64 years (73%), and was geographically concentrated in the Dakar (62%) and Thies (38%) regions. The overall SARS-CoV-2 positivity rate reflects the national testing strategy implemented during the main epidemic waves in Senegal [18–20]. To demonstrate the analytical utility of the standardized OMOP database, descriptive characterization was performed using the ATLAS platform. The following results illustrate the capacity of the OHDSI ecosystem to generate standardized indicators directly from the CDM without custom analytic pipelines.

Table 6 presents these positivity proportions by sociodemographic group. Because the testing population combined symptomatic referrals, contact tracing, traveler screening and community campaigns, these figures reflect the operational testing strategy and must not be interpreted as adjusted estimates of infection risk or population prevalence.

The prevalence of clinical symptoms was assessed using standardized condition and observation records. Fever was the most frequently reported symptom, affecting 6,233 individuals (47.6%), followed by headache in 3,412 individuals (26.1%). Soreness was observed in 650 individuals (5.0%), while pain and joint pain were each recorded in 266 individuals (2.0%). Muscle pain was reported in 166 individuals (1.3%). Gastrointestinal symptoms were less frequent, with vomiting in 131 individuals (1.0%) and diarrhea in 123 individuals (0.9%). For all listed symptoms, the average number of records per person was 1.0, indicating that symptoms were generally recorded once per individual.

Among the 13,090 individuals included in the standardized OMOP database, 52 deaths were recorded, corresponding to an overall case-fatality proportion of approximately 0.4%. Deaths occurred predominantly among middle-aged and older adults in both sexes. This low case-fatality proportion is consistent with the broad national testing strategy, which included travelers, community screening, and contacts in addition to symptomatic patients.

Together, these descriptive results confirm that the standardized OMOP database can be directly leveraged within the OHDSI analytical ecosystem to generate reproducible, standardized indicators.

## Discussion

This study documents the implementation of the OMOP Common Data Model for COVID-19 surveillance data in Senegal. By converting a large, heterogeneous, multi-source SARS-CoV-2 testing registry into a compliant OMOP CDM v5.4 database, we demonstrated that public health surveillance data collected under emergency conditions can be aligned with international interoperability standards in a low-resource, francophone African setting. Extending prior OMOP work in Africa [10,11,15–17], this implementation focuses specifically on pandemic surveillance data and addresses challenges particular to francophone contexts, including the translation of clinical terminology and the mapping of local drug brand names to international vocabularies. The ETL pipeline achieved complete transfer for eight of the eleven source-populated OMOP tables, including person, visit_occurrence, observation_period, measurement, death, location, care_site, and specimen. The two tables with incomplete transformation, such as condition_occurrence (95.3%) and observation (68.1%) reflect known limitations in vocabulary coverage for context-specific variables rather than systematic pipeline errors. The observation table gap was primarily driven by the 103 occupation categories in the source data, many of which lacked standardized equivalents in the OHDSI vocabulary. This challenge is consistent with findings from other African implementations: Ochola et al. reported that variables such as childhood vaccinations and socioeconomic status could not be mapped to existing OMOP concepts [15], while Yankam et al. encountered similar difficulties with local healthcare roles and profession-specific terms in Cameroon [17]. These gaps highlight the broader need for vocabulary extensions tailored to African public health contexts, an effort currently underway through the OHDSI Africa Chapter Vocabulary Working Group [10,11].

The corrected DQD pass rate of 98% indicates high structural and semantic integrity of the standardized database. This result can be contextualized against other published OMOP implementations in Africa and beyond. Ochola et al. reported a 98% overall pass rate for COVID-19 sero-surveillance data from the Nairobi Urban HDSS [15], while Yankam et al. achieved a 96% corrected pass rate for respiratory disease data at Douala General Hospital in Cameroon, which improved to 100% after iterative ETL verification [17]. In a North American setting, Marteau et al. evaluated the research data warehouse of a large multi-hospital pediatric health system (Shriners Children’s) and reported an overall pass rate of 83.2% across 1,125 DQD checks [29]. More broadly, the DQD has become the reference framework for OMOP data-quality assessment: Blacketer et al. formalized its conformance, completeness, and plausibility checks combining verification and validation [30], and the same approach was subsequently used to drive iterative, DQD-guided data-quality improvement across the federated databases of the European Health Data and Evidence Network (EHDEN) [31]. Our corrected pass rate of 98% is therefore consistent with - and at the upper end of - results reported for well-established OMOP implementations, suggesting that rigorous ETL design can achieve high data-quality standards even when working with operational surveillance data in resource-constrained environments. The detailed analysis of the 19 failed DQD checks showed that conformance and completeness failures were the most frequent categories (8 and 9 of 19, respectively), the conformance failures being dominated by foreign-key references that reflect the use of placeholder concept_id values (concept_id = 0) for metadata fields with no meaningful equivalent in surveillance data. This is a structural feature common to OMOP implementations based on non-clinical data sources [10,15,17] and does not inherently indicate data quality problems.

It is important to note, however, that the DQD evaluates structural conformance, completeness, and plausibility but does not verify the semantic correctness of source-to-concept mappings [21]. A high DQD pass rate therefore does not preclude residual mapping errors, particularly for locally specific or context-dependent terms. In our implementation, we addressed this limitation through systematic expert validation of all Usagi mappings, but we did not implement a formal dual-mapping procedure with inter-rater agreement assessment, which would further strengthen reliability.

The use of the ATLAS platform enabled rapid and reproducible descriptive analyses directly on the standardized OMOP database without custom analytic pipelines. Sociodemographic and clinical patterns observed in the standardized data were broadly consistent with published descriptions of COVID-19 in Senegal [18,19], providing indirect validation of the transformation. The clinical symptom profile, characterized by frequent fever and headache, aligns with international reports [32]. The low overall case-fatality proportion (0.4%) is consistent with the broad national testing strategy, which encompassed travelers, community screening, and contacts in addition to symptomatic patients.

The key value of these descriptive results lies not in their epidemiological novelty but in demonstrating that the same standardized queries and cohort definitions can now be applied across any OMOP-compliant database. This interoperability is the capability that pan-African and global initiatives such as INSPIRE seek to foster [10,11], and the existence of a high-quality OMOP CDM database is a prerequisite for meaningful participation in federated analyses.

Several practical lessons emerged from this implementation that may be useful for other teams, particularly in francophone African settings, undertaking similar OMOP standardization projects.

First, the translation of clinical data from French to English was not a one-time preprocessing step but an iterative process interleaved with vocabulary mapping. Initial translations that appeared linguistically correct often produced low Usagi match scores because the English terms used did not align with the specific terminology in OHDSI vocabularies. Revising translations to better approximate SNOMED CT or LOINC phrasing substantially improved mapping quality. Teams working with non-English source data should anticipate this iterative cycle and allocate time accordingly.

Second, among the OHDSI tools, WhiteRabbit and Rabbit-in-a-Hat were most valuable in the early design phase for understanding the source data structure and planning the mapping strategy. Usagi was essential but required substantial manual oversight - only 76.6% of mappings were approved after review and the remaining flagged mappings required clinical judgment. ATLAS proved immediately useful for validating the transformed data through visual exploration, often revealing mapping issues that were not apparent from DQD checks alone.

Third, local drug brand names common in Francophone Africa (Doliprane, Efferalgan, Perfalgan, Azicure) are generally absent from core RxNorm but available in RxNorm Extension. Teams should ensure they download and load the RxNorm Extension vocabulary from ATHENA in addition to the core vocabularies.

Fourth, the ETL pipeline required multiple iterations. Initial loads revealed referential integrity issues, unexpected NULL values, and mapping errors that were corrected through progressive refinement. Maintaining all ETL scripts under version control and generating detailed processing logs at each stage was critical for traceability and debugging.

The ETL pipeline and all associated scripts are publicly available and version-controlled, enabling future re-execution as new surveillance data become available. Sustaining this OMOP infrastructure will require institutional commitment to ongoing vocabulary updates, periodic ETL re-runs, and data governance oversight.

At IRESSEF, the data management team is positioned to maintain and extend this infrastructure as part of its broader digital health mandate.

Future work will focus on extending the OMOP infrastructure to other priority disease areas and data sources in Senegal, enhancing longitudinal linkages across health information systems, and establishing federated analysis workflows that preserve local data governance while enabling research collaboration. As OMOP vocabularies evolve and the French OMOP vocabulary extension matures, certain semantic mappings may need to be revisited to improve coverage of locally specific clinical terms.

Several limitations should be acknowledged. First, the source dataset was an operational surveillance registry, not a comprehensive clinical database. Testing strategies, underreporting of mild symptoms, and incomplete recording of comorbidities and treatments may introduce selection biases that limit the scope of downstream analyses. Second, the analyses presented are intentionally descriptive; we did not perform predictive modeling, causal inference, or comparative effectiveness studies, and the standardized database should not be considered sufficient for such purposes without additional data sources and dedicated study designs. Third, semantic mappings were validated by clinical and data science experts, but a formal dual-mapping procedure with inter-rater agreement assessment was not implemented. Fourth, the observation table achieved only 68.1% transformation completeness, reflecting vocabulary gaps for occupation categories and contextual variables that constrain certain analyses. Finally, the observation_period table was structured with one record per visit rather than one per person, which deviates from the standard OMOP convention and should be considered when designing longitudinal analyses on this database.

## Conclusion

This study demonstrates the successful standardization of COVID-19 surveillance data from Senegal into OMOP CDM v5.4, achieving a corrected DQD pass rate of 98% and complete transformation for most domain tables. Using open-source OHDSI tools and a documented ETL pipeline, we transformed heterogeneous, real-world surveillance data into an interoperable database that can be leveraged within the OHDSI analytical ecosystem. The scope of the findings is conditioned by the characteristics of the underlying surveillance data, which provide limited information on comorbidities, care pathways, and long-term outcomes. Adaptation to other diseases and settings will require accounting for local information systems, coding practices, and governance frameworks. Beyond its technical contribution, this work establishes a reusable digital health infrastructure that can support outbreak preparedness, observational research, and participation of Senegal in global federated health data networks. While the current surveillance dataset alone is not sufficient for predictive modeling, causal inference, or risk estimation, the OMOP infrastructure established here provides the foundation for such analyses once enriched with complementary clinical data sources or integrated into multi-site federated studies. By showing that international data standards can be effectively implemented in a low-resource, francophone public health setting, this study contributes to reducing the underrepresentation of African populations in global real-world data research.

## Data Availability

All ETL materials, vocabulary mapping specifications, SQL scripts, and data quality assessment outputs underlying this study are publicly available at https://github.com/odiop88/omop-covid19-iressef and are archived with a persistent identifier at https://doi.org/10.5281/zenodo.20218885. The underlying individual-level surveillance data cannot be shared publicly because they contain sensitive personal health information whose dissemination is restricted under Senegalese Law No. 2008-12 of 25 January 2008 on the Protection of Personal Data and under the data governance policy of IRESSEF. De-identified data may be made available to qualified researchers for legitimate scientific purposes upon reasonable request and subject to a data-sharing agreement requests should be addressed to the Data Management Department of IRESSEF, Dakar, Senegal.

https://github.com/odiop88/omop-covid19-iressef/tree/master

## Acknowledgment

The team acknowledges with gratitude the support of their respective institutions for the production of this manuscript.

## Ethics statement

This study is a secondary analysis of fully anonymized COVID-19 surveillance data that were routinely collected by IRESSEF and partner sites during the national public health response to the SARS-CoV-2 pandemic. No new data were collected for this study, and no identifiable personal information was used: all direct and indirect identifiers were removed and replaced with anonymized surrogate identifiers before any analysis, in accordance with Senegalese Law No. 2008-12 of 25 January 2008 on the Protection of Personal Data. Because the work involved only the methodological transformation of de-identified, routinely collected surveillance data, it did not constitute human-subjects research requiring prospective ethical review, and no individual informed consent was required for this secondary use. Data access and processing were carried out under the data governance framework of the Data Management Department of IRESSEF. The study was conducted in accordance with the principles of the Declaration of Helsinki.

## Supporting Information

**S1 Table. Source data dictionary**

**S2 File. STROBE checklist**

## Notes

### Competing Interest Statement

The authors have declared no competing interest.

### Funding Statement

The author(s) received no specific funding for this work.

